# Unraveling the “indirect effects” of interventions against malaria endemicity: A systematic scoping review

**DOI:** 10.1101/2024.05.08.24307059

**Authors:** Yura K. Ko, Wataru Kagaya, Chim W. Chan, Mariko Kanamori, Samuel M. Mbugua, Alex K. Rotich, Bernard N. Kanoi, Mtakai Ngara, Jesse Gitaka, Akira Kaneko

## Abstract

There is an urgent need to maximize the effectiveness of existing malaria interventions and optimize the deployment of novel countermeasures. When assessing the effects of interventions against malaria, it is imperative to consider the interdependence of people and the resulting indirect effects, without which the impact on health outcomes and their cost-effectiveness may be miscalculated. Here, we conducted a scoping review of existing literature on the indirect effects of malaria interventions. We observed a recent increase in both the number of reports and the variety of terms used to denote indirect effects. We further classified eight categories of comparative analysis to identify the indirect effects, proposed common terms for the indirect effects, and highlighted the potential benefits of mathematical models in estimating indirect effects. Improving the study design and reporting the indirect effects of malaria interventions will lead to better informed decisions by policymakers.

## Introduction

The global fight against malaria has become increasingly challenging in recent years. Despite concerted scale-up of intervention tools, such as the widespread distribution of long-lasting insecticidal nets (LLINs), rapid diagnostic tests (RDTs), and artemisinin-based combination therapies (ACTs), the estimated global case incidence of malaria in the past few years has stagnated at around 58 cases per 1,000 population at risk, while the global mortality rate has remained at approximately 14 per 100,000 population at risk^1^. Moreover, although malaria remains a leading cause of healthcare spending in endemic countries^2^, the amount invested in 2022 fell short of the estimated USD 7.8 billion required globally to achieve the Global Technical Strategy (GTS) targets set by the World Health Organization (WHO)^1^. It is anticipated that high-income nations and other international funders will continue to prioritize their efforts to address emerging diseases such as COVID-19 through 2024^3^. In this context, there is an urgent need to re-evaluate existing malaria interventions for more effective deployment, along with the employment of novel countermeasures to reduce the malaria burden more efficiently and cost-effectively.

Malaria is a vector-borne disease transmitted by *Anopheles* mosquitoes. When measuring the effects of interventions against such diseases, it is crucial to consider the interdependence in people, often referred to as “dependent happenings”^4^. For instance, in malaria-endemic settings, a decline in the number of malaria-infected individuals or mosquitoes will reduce parasite reservoirs and means of transmission in a community, leading a lower possibility of infection among all community members. Consequently, malaria control measures implemented in a community are expected to yield direct benefits for individuals receiving the interventions and indirect benefits for both individuals receiving and not receiving the interventions. Indirect effects can be defined as the unintended positive or negative consequences of an intervention that influences disease transmission or health outcomes. Thus, without proper consideration of the indirect effects, malaria interventions’ impacts on health outcomes and their cost-effectiveness may be overestimated or underestimated. Therefore, adopting a comprehensive and standardized approach to identify both direct and indirect effects is imperative to gain a detailed understanding of intervention impacts. Moreover, evidence of indirect effects will influence policy makers’ decision making. If the direct effects are equivalent, an intervention that broadly benefits those who do not receive the intervention is preferable to one that benefits only a limited number of people who receive the intervention.

The concept of indirect effects of malaria intervention, especially LLINs, has long been well known. Nevertheless, the description of indirect effects in the WHO guidelines for vector-borne mosquito control only briefly states that community-level effects of ITNs have not always been observed^5^. In addition, the scientific literature on malaria interventions that explicitly differentiate and thoroughly analyze their indirect effects is currently limited^6^. A recent systematic review of the indirect effects of interventions on health in low- and middle-income countries by Benjamin-Chung et al^7^. included only two malaria-related studies. Moreover, the methodology of measuring the indirect effects greatly varies, and the terms indicating the indirect effects are not standardized (e.g., community effects, spillover effects, mass effects, herd effects, area-wide effects, spatial effects, and positive externalities). To address these knowledge gaps, we conducted a scoping review to summarize how the indirect effects of malaria interventions were analyzed and reported.

## Methods

### Search strategy and selection criteria

We followed the recommendations of the Preferred Reporting Items for Systematic Reviews and Meta-Analysis extension for scoping reviews (PRISMA-ScR)^8^. The study protocol is available at elsewhere (https://inplasy.com/inplasy-2023-6-0025/).

#### Literature search

We searched PubMed, Web of Science, and EMBASE by title and abstracts. In addition, for grey literature, we searched OAIster by keywords. Searches were conducted in June 2023. We set the search terms as follows: (“malaria” OR “plasmodium”) AND (“indirect effect*” OR “indirect protection” OR “herd effect*” OR “herd protection” OR “community effect*” OR “communal effect*” OR “community-level effect*” OR “community protection” OR “communal protection” OR “community-level protection” OR “peer effect*” OR “peer influence effect*” OR “mass effect*” OR “assembly effect*” OR “spillover effect*” OR “contextual effect*” OR “free-rider” OR “free rider” OR “free-riding” OR “free riding” OR “positive externality” OR “positive externalities” OR “dependent happenings”)

#### Eligibility criteria

We included studies that were conducted to quantify the indirect effects of any interventions for all species of malaria infection. We excluded non-original papers such as opinions and editorials. We only targeted articles written in English. We defined indirect effects as the impact accrued by either the non-intervention or intervention group, stemming from alterations in malaria parasite or mosquito populations within a community consequent to an intervention. It should be noted that simple group comparisons between treatment and control (or baseline) groups/clusters are regarded as total effects. Studies that reported only total effects were excluded from our review. However, if the treatment coverage in the community was considerably low, the group comparisons between treatment and control were considered indirect effects and were included in our review.

#### Study selection

We imported the data for each relevant publication into reference software (Rayyan, https://www.rayyan.ai/). Prior to the initial screening, duplicate records were deleted automatically. In the first review step, two reviewers (YKK, SMM) screened all records by title and abstract according to the eligibility criteria. Any discrepancies in the process were resolved by discussion between both reviewers. Once a record was selected, its full text was reviewed by at least two of five reviewers (YKK, WK, CWC, MK, and AKR). Specific data (see the section “*Data extraction and analysis*”) were recorded and summarized in a tabular form through this second review step. Any disagreement was addressed through discussion. Additional reference and citation searches were also conducted. The reference lists of the articles identified during the search were scanned manually, and eligible articles were included in the full-text reading.

#### Data extraction and analysis

We used a standardized data collection form that follows the PRISMA guidelines for scoping reviews^8^ to obtain the following information from each record: title, name of authors, year of publication, region, country, study type, malaria parasite species, type of interventions, type of outcomes, separate estimated indirect effect for different conditions (yes/no), pre-specified to measure indirect effect (yes/no), secondary analysis of previous study (yes/no), methods of indirect effects estimation, terms of indirect effects, and if positive or negative indirect effects observed (yes/no). A detailed description of the extracted data is in Supplementary Table 1. Standardized labels were made for each term for inconsistencies of words, as listed in Supplementary Table 2.

#### Quality of study methodology for estimating indirect effect

We utilized the classification of risk of bias for indirect effect estimation proposed by Benjamin-Chung et al^7^. We only assessed the risk of bias for field epidemiological studies, excluding mathematical modeling studies and experimental hut trials. Each eligible study was classified as “very low”, “low”, “medium”, or “high” in terms of the reliability of indirect effects estimation.

## Results

### Study selection

Figure 1 illustrates a PRISMA flow diagram depicting the identification, screening, eligibility, and exclusion process of the studies. A total of 664 articles were identified through database searches (n = 570) and other sources (n = 94). Three hundred sixty-eight duplicate articles were removed. Thirty-eight articles met the eligibility criteria after review of titles and abstracts; 258 studies were excluded for one or more of the four following reasons: 1) different meanings of indirect effect, 2) not malaria-specific intervention, 3) not intervention study, and 4) not reporting indirect effect. Notably, among the studies excluded because of different meanings of indirect effect, 14 studies evaluated the indirect relationship between COVID-19 and malaria^9–22^, and one study was a causal mediation analysis^23^. Six articles were added from a manual search of reference lists of the 38 eligible articles from the initial screening. Of these 44 studies, 31 were included in this review after full-text reading. The reasons for exclusion in the full-text reading were 1) reporting total effect only (n = 7), 2) opinion or review article (n = 3), 3) estimating indirect effect in the context of mediation analysis (n = 2), and 4) not reporting indirect effect (n = 1).

**Figure 1:**
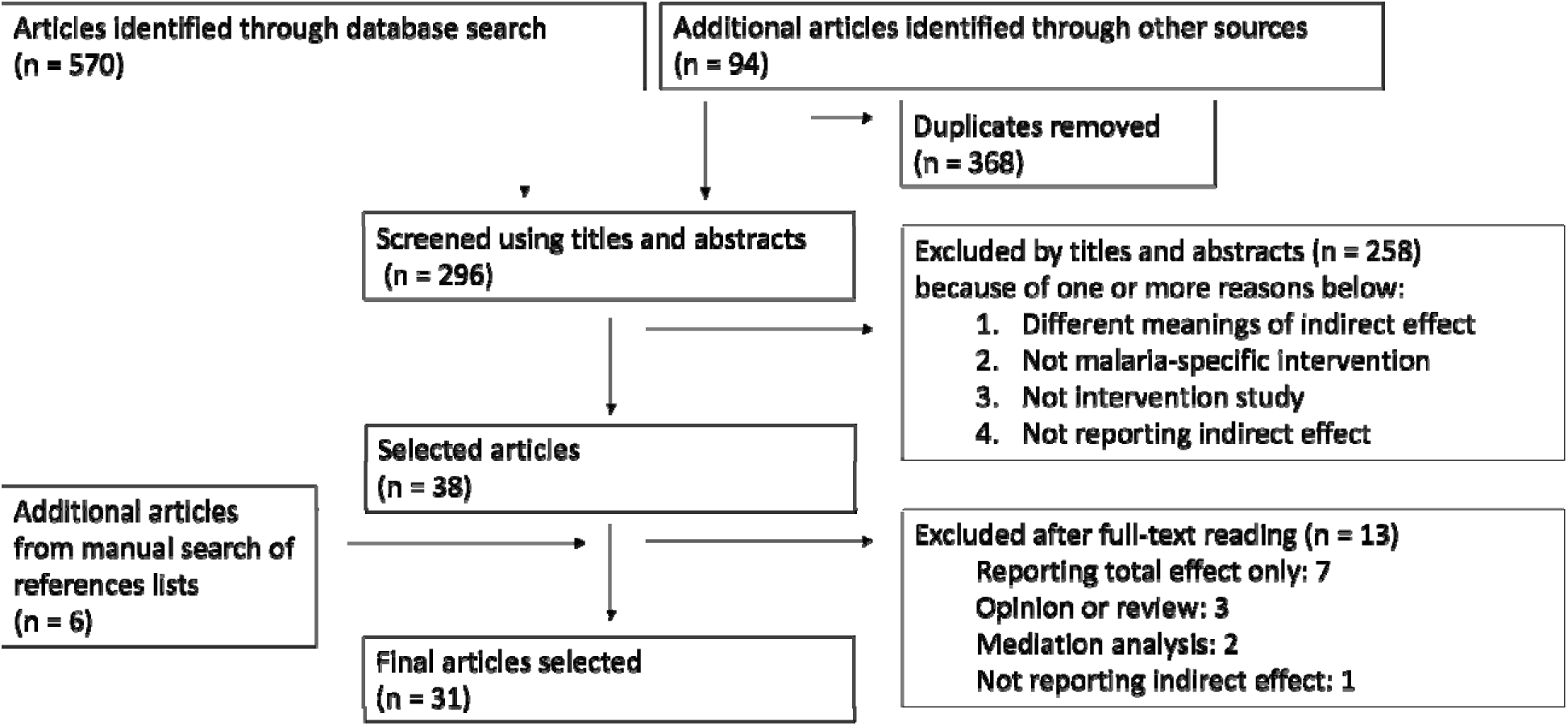
PRISMA flowchart of study selection

### Study characteristics

Details of the 31 reviewed studies are summarized in Table 1. Most studies were set in African countries (n = 24; 77%) and examined the indirect effects of interventions on *P. falciparum* (n = 18; 58%). Temporal trends in study types, intervention types, and terms used to describe indirect effects are illustrated in Figure 2. Overall, until year 2000, very few studies purposefully reported indirect effects. Subsequently, there was a sharp increase in reporting from 2001 to 2005, followed by a gradual decline. Since 2016, there has been an upward trend once again (Figure 2a). The most common study type was mathematical modeling (n = 9; 29%), followed by cross-sectional surveys (n = 6; 19%) and re-analysis of cluster-randomized trials (n = 6; 19%) (Figure 2a). The most common interventions were insecticide-treated nets (ITNs) or LLINs (n = 17; 55 %). Until 2015, the focus was primarily on ITN/LLIN-related interventions. However, since 2016, reports on various interventions such as house modification, intermittent preventive treatment (IPT), seasonal malaria chemoprevention (SMC), and mass drug administration (MDA) have emerged. (Figure 2b) The most common terms used for indirect effects were “communal” or “community” effect/benefit/protection (n = 23; 74%), followed by “mass” or “mass killing” effect/benefit/protection (n =11; 36%). Until 2015, the use of communal/community effect and the mass effect dominated, but more recently, various terms have come into use, including herd effect, indirect effect, spatial effect, and spillover effect (Figure 2c). Among 21 studies eligible for quality assessment of evidence, 6 (29%) had high-quality evidence, 7 (33%) had moderate, 5 (24%) had low, and 3 (14%) had very low-quality evidence. Of studies with high-quality evidence, 5 (83%) used cluster-randomized designs.

**Figure 2:**
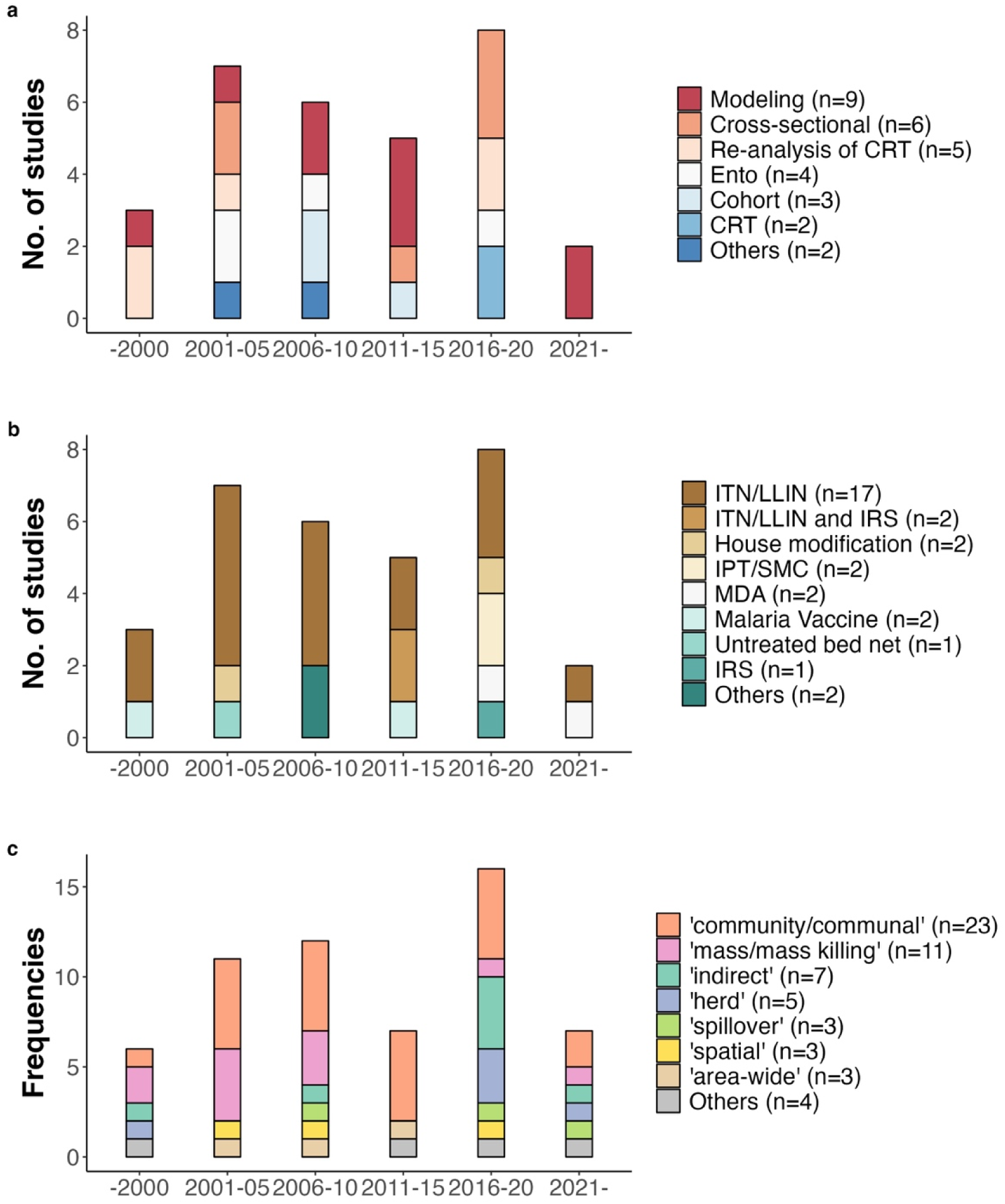
Time trend of study characteristics; a) study type, b) intervention type, c) term used to describe the concept of indirect effects. Note that for c), the total number of terms in the graph does not correspond to the total number of studies (n=31), as multiple terms can be used in a single paper. CRT: cluster randomized trial, Ento: entomological survey, ITN: insecticide-treated net, LLIN: long-lasting insecticide-treated net, IRS: indoor residual spray, IPT: intermittent preventive treatment, MDA: mass drug administration. For the study type, “Others” included analysis of passive case detection using surveillance data. For intervention type, “Others” included access to free antimalarials and target subsidies of ITNs. Regarding indirect effects terminology, “Others” included assembly effects, population effects, group-level effects, positive externality, and dependent happenings.

**Table 1:**
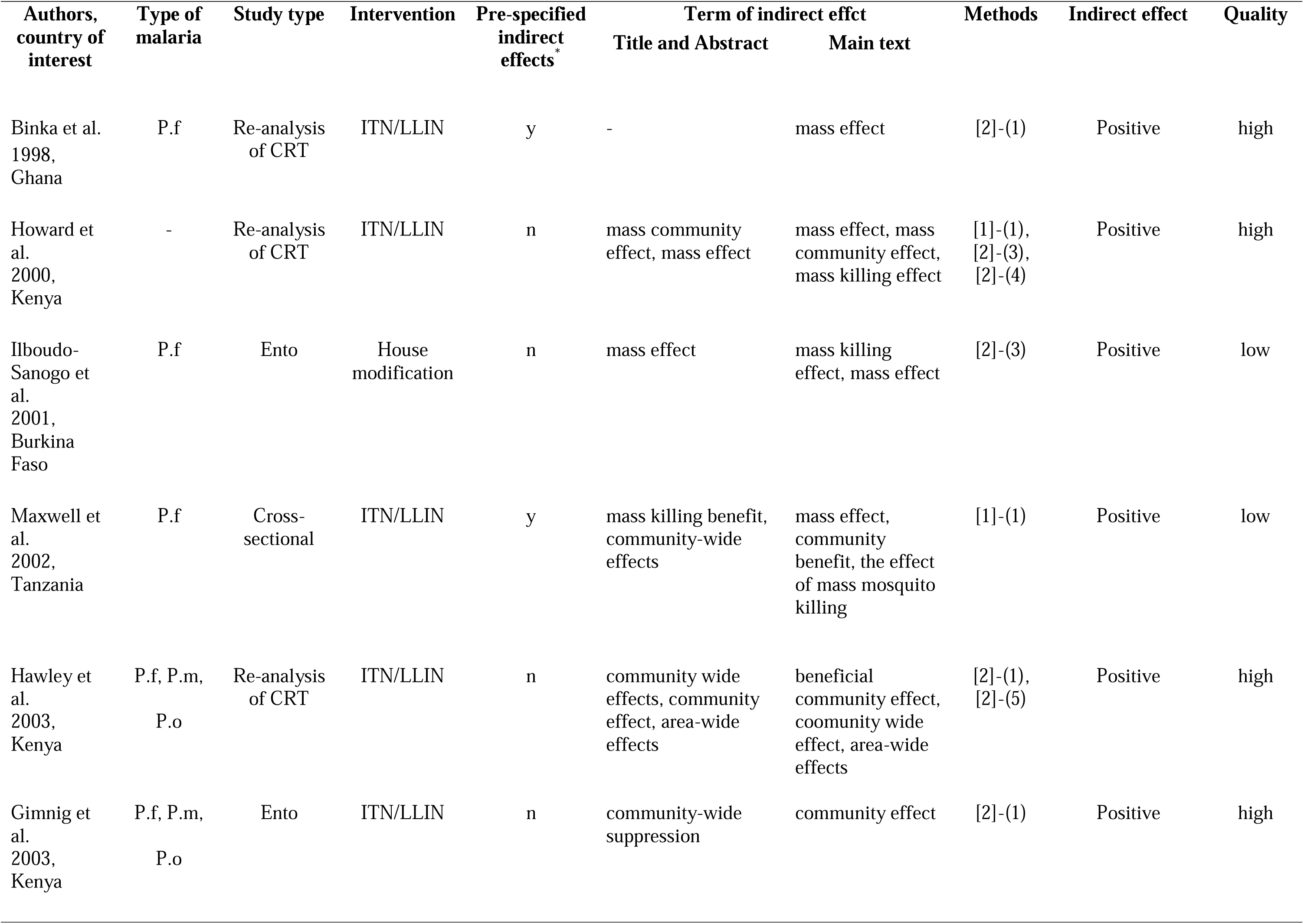
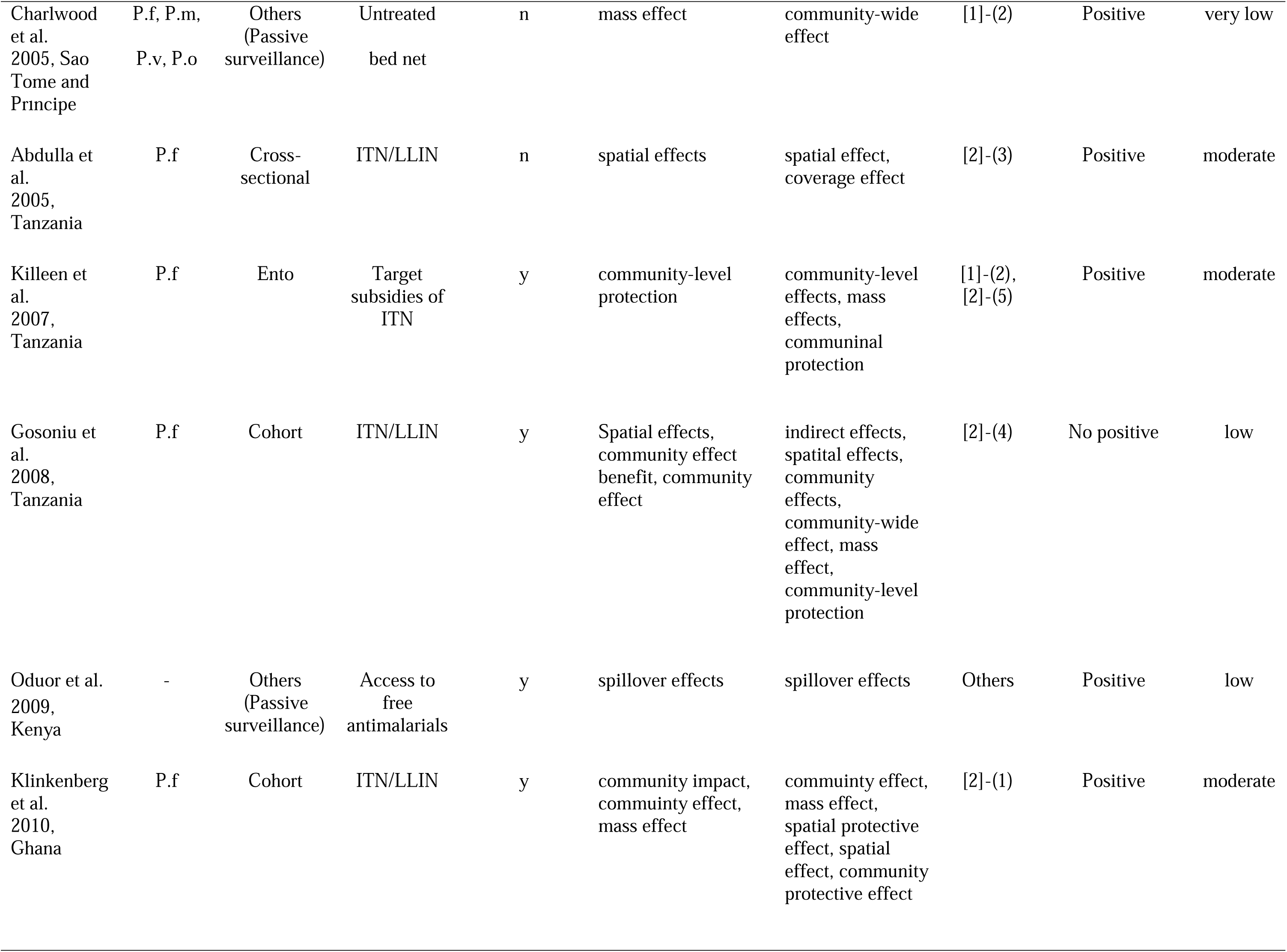
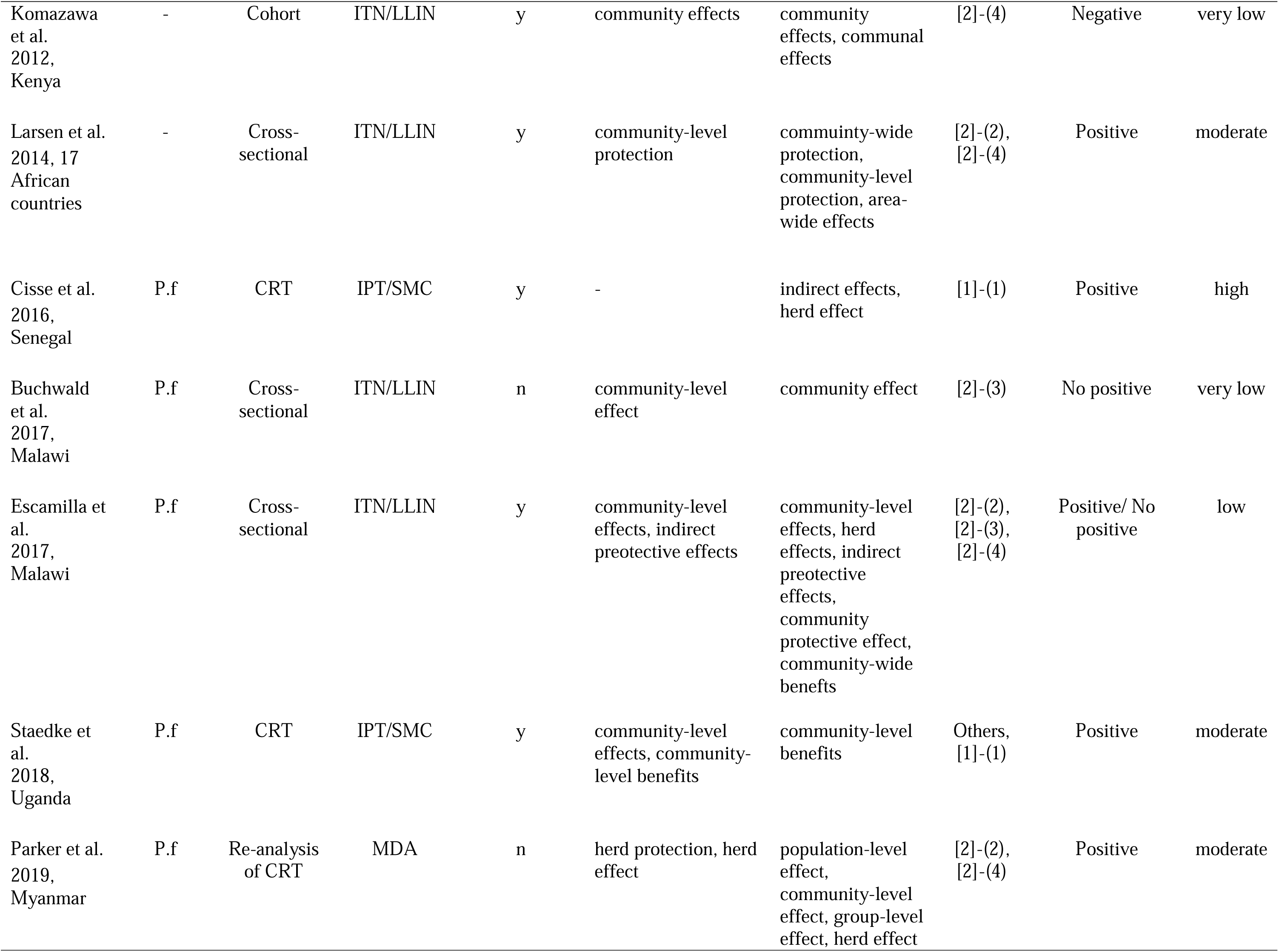
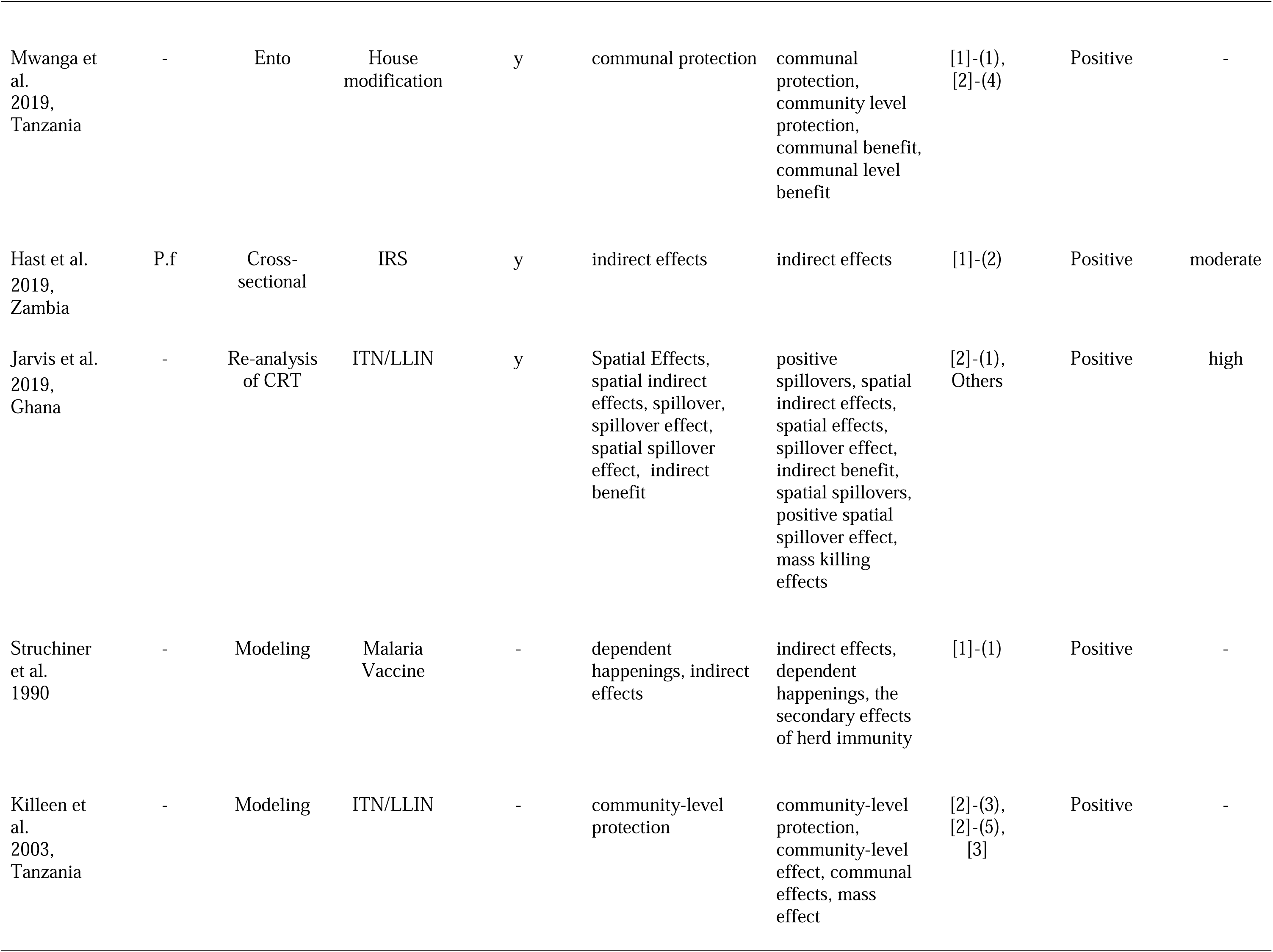
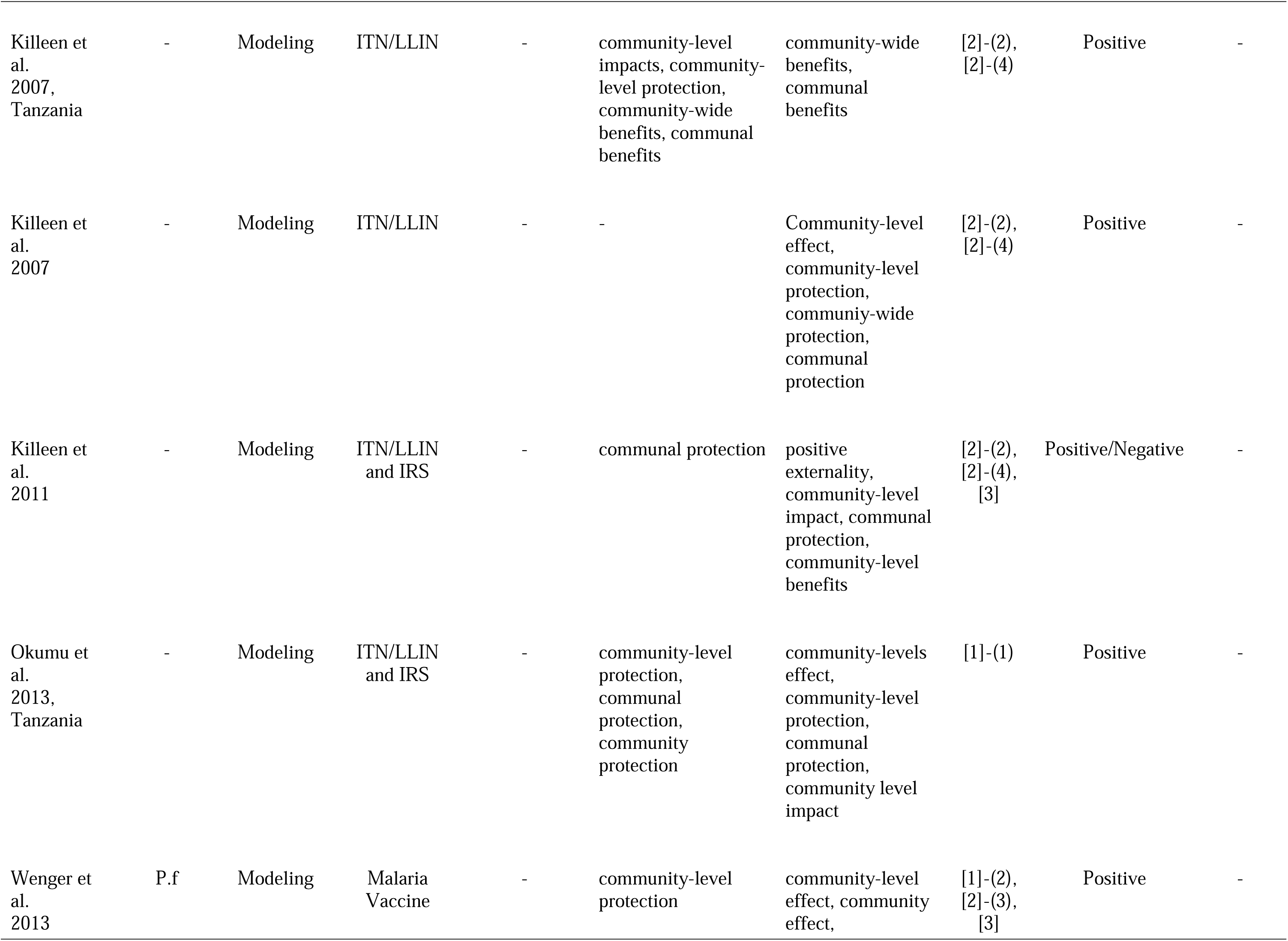
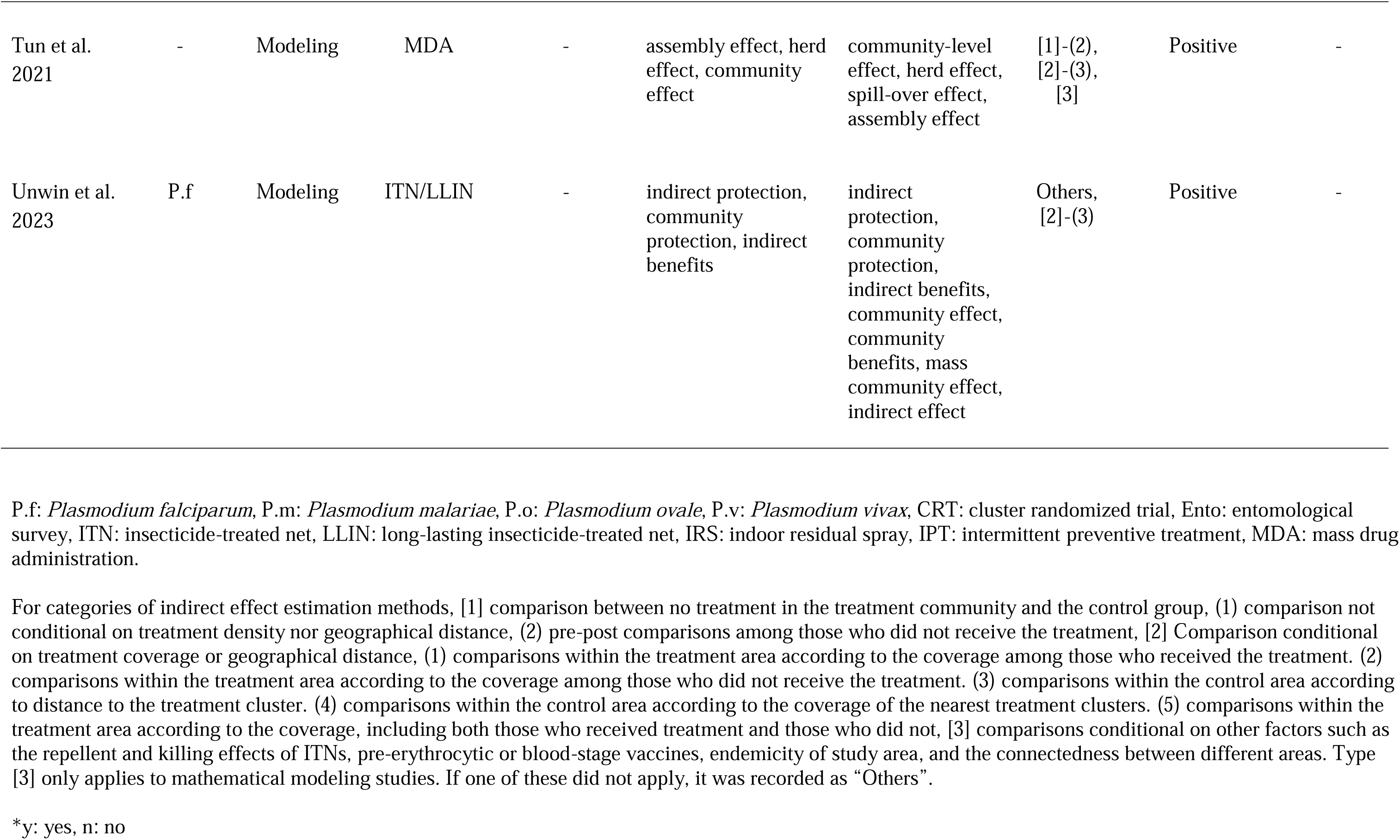
Characteristics of 31 included studies.

### Overview of methods for indirect effect analysis

Among all included studies, each intervention’s indirect effect was evaluated in relation to reductions in malaria transmission. Figure 3 shows the categories of methods for indirect effect estimation identified through this review. In addition, a detailed description of the methods by intervention type is listed in Supplementary Table 3.

**Figure 3:**
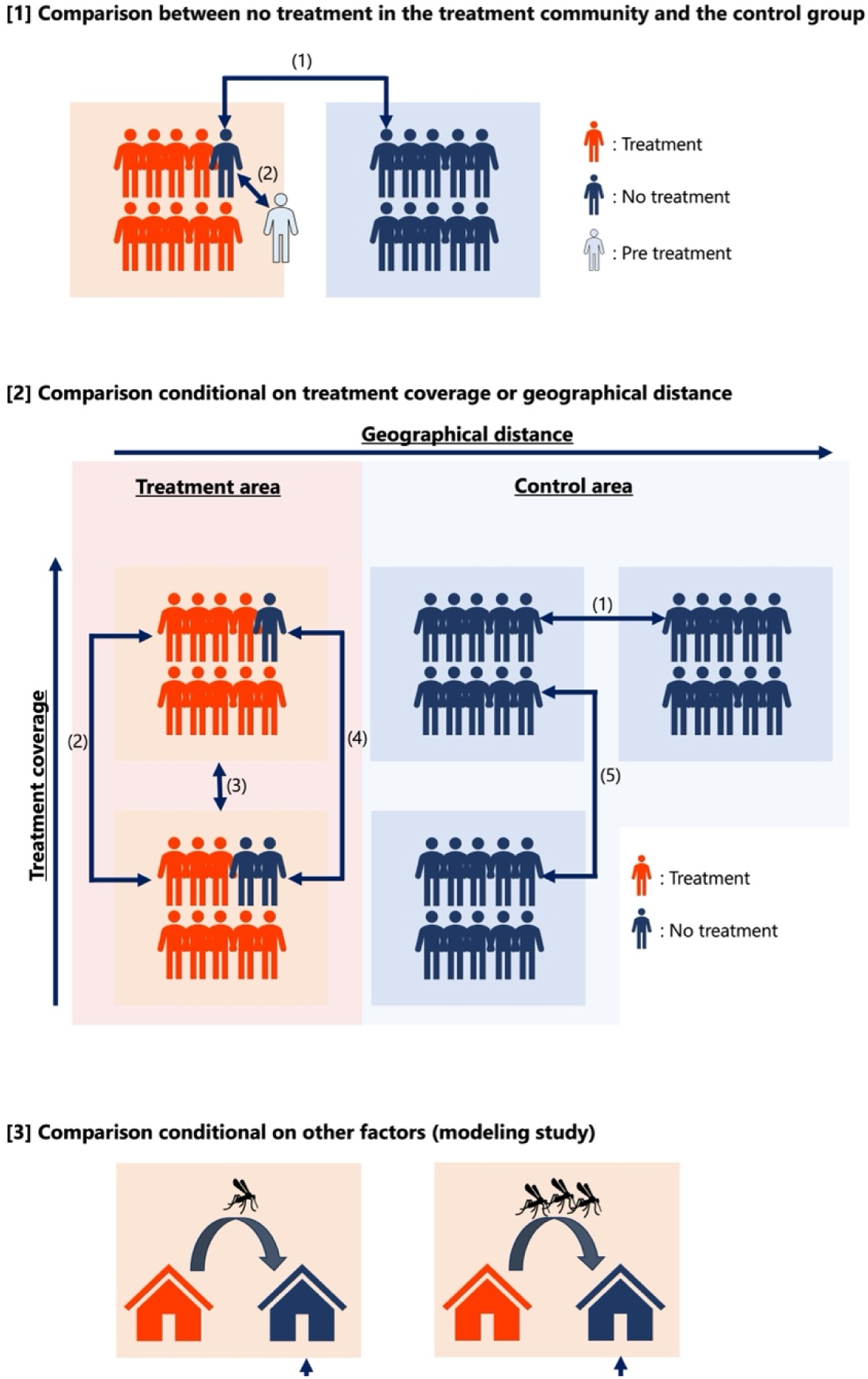
Categories of indirect effect analysis methods. [1] comparison between no treatment in the treatment community and the control group, (1) comparison not conditional on treatment density nor geographical distance, (2) pre-post comparisons among those who did not receive the treatment, [2] Comparison conditional on treatment coverage or geographical distance, (1) comparisons within the control area according to distance to the treatment cluster. (2) comparisons within the treatment area according to the coverage among those who received the treatment. (3) comparisons within the treatment area according to the coverage, including both those who received treatment and those who did not. (4) comparisons within the treatment area according to the coverage among those who did not receive the treatment. (5) comparisons within the control area according to the coverage of the nearest treatment clusters, [3] comparisons conditional on other factors such as the repellent and killing effects of ITNs, pre-erythrocytic or blood-stage vaccine efficacy, endemicity of study area, and the connectedness between different areas. Type [3] only applies to mathematical modeling studies. If one of these did not apply, it was recorded as “Others”.

### Field studies (epidemiological and entomological studies)

Among field studies, including epidemiological and entomological studies, 59% pre-specified analysis of indirect effects (n = 13). Comparisons of non-treatment populations in intervention communities with non-intervention communities or pre-post analyses of these populations ([1]-(1) and [1]-(2), respectively, in Figure 3) were employed by eight studies^24–31^. On the other hand, comparison among no-intervention individuals/groups according to distance to the treatment household or the treatment coverage within a certain distance range were employed by 16 studies ([2] in Figure 3)^24,27,30,32–44^.

Comparisons conditioned on the distance to nearest intervention were reported in five studies^32,34,35,38,44^ ([2]-(1) in Figure 3), all of which evaluated the impacts of ITN. There were two analytical approaches. One was to compare between groups stratified by the distance category set at 100 – 400 m intervals, with the most distant group as the reference. In all studies, households without ITNs within 300 – 400 m of households with ITNs had the lowest risk of malaria-related outcomes (e.g. malaria parasitemia, mosquito density, anemia, and all-cause mortality). Another approach to measuring indirect effects by conditioning on distance was trend analysis, in which regression was performed with distance as an explanatory variable. Around year 2000, researchers simply incorporated distance into the model as a continuous variable^32,35^, but recently, Jarvis et al. have used a quadratic term to account for the nonlinearity called “distance decay” in spatial analysis^44^. The study reported that for every additional 100 m that a control household was from an intervention household, the all-cause mortality for children aged 6–59 months increased by 1.7%^44^.

Regarding interventions conditional on treatment coverage, two patterns were observed: comparing among intervention populations ([2]-(2) and [2]-(3) in Figure 3) and among non-intervention populations ([2]-(4) and [2]-(5) in Figure 3). The definition of the areal unit for calculating intervention coverage varied from study to study, with a single cutoff determined by a 100-m to 3-km radius of the subject’s household^34,36,42,43^, multiple distances used in an exploratory manner^24,37,39^, and using primary sampling units^40,41^. There were also two approaches to analyzing indirect effects: one in which groups were stratified by intervention coverage and the other in which regression analysis was performed by incorporating intervention coverage as an explanatory variable.

Several approaches other than the above-mentioned methodology were used to evaluate the indirect effects (categorized as “Others” in Table 1). Jarvis et al. (2019) showed that the treatment effects changed after reallocating the treatment and control cluster assignments based on the distance to the nearest treatment cluster^44^. Oduor et al. (2009) suggested positive indirect effects by confirming that the direct treatment effects were enhanced when spillovers to the neighboring sub-locations were accounted for^45^. In addition, Staedke et al. (2018) evaluated the effect of IPT in school children by comparing the reduction in malaria prevalence in all age groups between the intervention and control clusters^29^. The risk reduction was regarded as a community-level effect because the treatment coverage was considerably low (only school children among all age groups).

Only two studies examined indirect effect heterogeneity^40,42^. Escamilla et al. (2017) reported that an increase in community bed net coverage was significantly associated with a decrease in malaria prevalence among children under five years and 5 – 19 year-olds, but no association was observed among adults older than 20 years^42^. In another study by Larsen et al. (2014), subgroup analyses were performed, stratified by rural versus urban areas and low versus high malaria transmission; however, no significant effect heterogeneity was observed^40^. In four studies, positive indirect effects were not observed, or negative indirect effects were observed with increased treatment coverage^37,39,41,42^. All four studies were observational studies. Among the field studies, 59% pre-specified analysis of indirect effects (n = 13).

### Mathematical modeling studies

Among nine studies employing mathematical models^46–54^, two-thirds (n = 6) aimed to estimate the indirect effects of ITNs/LLINs, comparing outcomes before and after the intervention in the non-intervention group or altering parameters of intervention coverage through simulation. No mathematical modeling studies conducted a comparison based on distance conditioning, likely due to the infrequent use of spatial data in malaria transmission models. One notable characteristic of mathematical models is their ability to vary efficacy by changing more detailed parameters of interventions, such as the repellent and killing effects of ITNs^50^, vaccine target for pre-erythrocytic or blood-stage *P. falciparum*^52^, endemicity of study area^53^, and the connectedness between different areas^47,53^ ([3] in Figure 3).

Another distinctive method for estimating indirect effects involves using counterfactual hypothetical models. Unwin et al. (2023) disentangled the direct and indirect effects of ITNs^54^ by maintaining the entomological inoculation rate (EIR) over time in certain scenarios, thereby breaking the link between current malaria endemicity and the human force of infection.

## Discussion

To our knowledge, this is the first systematic scoping review on the indirect effects of malaria intervention. We reviewed studies whose titles or abstracts included terms indicative of indirect effects (except some articles from manual searches) and revealed that the number of such studies has increased in recent years, especially for interventions other than ITNs/LLINs. In addition, although not included in this review, an opinion piece^6^ and a methodology study^55^ have recently been published relating to the indirect effects of malaria intervention. Most recently, in 2023, a study intended to estimate both indirect and direct effects of reactive, focal chemoprevention and vector control interventions was made available as a preprint^56^. In light of the increasing interest in the indirect effects of malaria interventions, a scoping review summarizing previous studies is pertinent and salient.

Several terms have been used to convey indirect effects. Apart from the “mass/mass killing” effect, which refers to the reduction of malaria transmission by decreasing the mosquito abundance or density through insecticides, other terms such as community effects, spillover effects, mass effects, and herd effects have been used interchangeably to denote indirect effects. Historically, indirect effects of malaria control interventions have often been labeled as community effects, especially for ITNs/LLINs (Supplementary Figure 1) and in the WHO vector control guideline^5^. In recent years, there has been more diversity in the terminology, particularly for interventions other than ITNs/LLINs. This diversity of terminology may create confusion and make it difficult for literature search on this topic. We propose using either community effects or spillover effects, a widely used term in general epidemiology^7,57^, when reporting indirect effects in malaria control, regardless of the type of intervention.

We found that studies varied in their methodology for estimating indirect effects, although most can be typified into eight categories (Figure 3). Since malaria parasites are transmitted via mosquitoes, it is appropriate to make comparisons conditional on distance to account for mosquito flight range or intervention density within that range. Comparisons between non-treatment groups conditional on distance from the treatment group were only conducted in studies on vector control such as ITNs/LLINs, while studies on interventions against parasites such as MDA, IPT/SMC, and vaccine were conditioned by treatment coverage (Supplementary Table 3). Future studies investigating the effectiveness of malaria interventions could draw on these methods, taking into account geographical characteristics and the feasibility of each study.

When comparing the non-treated within intervention clusters, double-randomized trials^58^, which allow for the strongest inference by minimizing selection bias and unmeasured confounding, are considered the recommended approach^7,57^. However, we did not find any studies in our review that performed two-stage randomization. One possible reason is that a double-randomized trial is not always feasible, especially in the evaluation of malaria interventions. Because of the additional allocation of controls within the intervention cluster, more samples or reduced intervention coverage are needed to obtain sufficient power for the estimation of the indirect effect. In addition, in malaria, there are interventions that target subpopulations in the community, such as IPT, SMC, and vaccination targeting children or pregnant women. In these interventions, untargeted individuals in the treatment group and their counterparts in the control group (i.e., individuals who would be ineligible if they were assigned to the treatment group) may be comparable, effectively emulating a cluster-randomized trial design, which would not necessarily require a two-stage randomization. If using a cluster-randomized design or analyzing observational studies in which ineligible populations are not comparable to eligible populations, matching should be considered. It should be noted, however, that even with matching, unmeasured confounding may remain, and external validity may be reduced^57,59^.

Other than changes in the number of malaria-infected individuals (drug or vaccine administration) or mosquitoes (vector control), indirect effects of interventions can manifest in two ways^7^: 1) individuals change their behavior because of the intervention and, in turn, influence the behavior of non-recipients in neighbors (social proximity), and 2) if a household member receives additional resources through the intervention, other household members will benefit because additional resources are available to the household (substitution). These indirect effects may not be trivial, and their relative magnitude may vary from setting to setting, which would necessitate intervention deployment plans tailor-made to suit area specificity, a lesson learned from the first Global Malaria Eradication Programme. We did not find studies reporting the indirect effects through these mechanisms that met our inclusion criteria. We excluded one study estimating the association between the proportion of nearby households receiving ITN subsidies and the probability of ITN use^60^ because net usage was the only outcome reported. Future research on the impact of changes in individual behavior through programs such as conditional cash transfers^61^ and subsidies based on malaria infection, morbidity, and mortality, especially when implemented alongside other malaria interventions, is warranted.

Four studies either did not identify a positive indirect effect or reported a negative indirect effect^37,39,41,42^. There are several reasons for not observing positive indirect effects. First, indirect effects, in general, tend to be smaller than direct effects, studies designed to detect direct effects as primary objectives are often underpowered to detect indirect effects^7^. For instance, vector control measures reduce malaria transmission by reducing EIR in the community, but EIR and parasite prevalence are not linearly related^62^, and a substantial EIR reduction would be required to reduce malaria prevalence among non-recipients.

Second, there is the potential confounder of residents’ behavior associated with both intervention compliance and the outcome. Residents’ compliance with interventions may depend on their perception of the risk of malaria transmission in the community and mosquito density^63^. For example, increasing community net usage is often associated with increasing mosquitos and malaria risk^64^. So, comparisons between non-recipients, especially when conditioned on coverage, may underestimate indirect effects. In addition, characteristics of non-recipients such as socio-economic status, healthcare access, and malaria preventive behavior may be different according to community treatment coverage, especially in an observational study setting^65^. Third, migration of infected individuals and mosquitoes between targeted and untargeted areas may have reduced the impact of the intervention in targeted areas^31^. No field studies conducted to date have taken into account these human and mosquito mobility to estimate indirect effects.

Recently, there has been a substantial upsurge in the number of mathematical modeling studies on malaria^66^. In agent-based models, estimating the impact of an intervention in the non-intervention population is straightforward within any simulation, thus we had expected a greater number of modeling studies that estimated indirect effects. However, only nine mathematical modeling studies were included in our review. It is possible that our screening, based on keywords in titles and abstracts, excluded many of these studies. This also supports the importance of our proposal on standardizing the terms used to refer to an indirect effect. An advantage of mathematical modeling is the ability to examine changes in indirect effects not only by varying the coverage of the intervention but also by adjusting other parameters, such as deterrent and insecticidal effects in the case of ITNs/LLINs, simultaneously. It would be beneficial to take advantage of mathematical models and consider parameters for which data are not reliably quantified. For example, the main advantage of house modification is that once installed, it remains semi-permanent. Therefore, its effect is less susceptible to variations in human behavior^67^, such as repurposing and inconsistent uses of LLINs^68^. Incorporating such behaviors into the model and estimating the indirect effects on those who do not receive the intervention will have important implications for the widespread implementation of the intervention.

One limitation of our study is that the search strategy may not have captured all relevant articles. We searched for keywords in the titles and abstracts, potentially missing studies that only reported the indirect effects of malaria interventions within the full text of the article. While efforts were made to manually include references cited for indirect effects, they were unlikely to be complete. Additionally, Benjamin-Chung et al. noted evidence of publication bias reporting for indirect effects^7^. Nonetheless, this review aimed to pave the way for improved design and reporting of future research on the indirect effects of malaria interventions. By highlighting this critical area, we hope to contribute to a more appropriate evaluation of intervention effectiveness.

In conclusion, our review notes an increase in the number of studies that measured the indirect effects of malaria interventions in recent years. We outline eight comparative schemes by which indirect effects of malaria interventions can potentially be quantified, and propose standardized terms for describing indirect effects. We further support the use of mathematical models to inform the evaluation of indirect effects of malaria interventions. Incorporating assessment of indirect effects in future trials and studies may provide insights to optimize the deployment of existing and new interventions, a critical pillar in the current fight against malaria globally. In addition, evidence about the cost-effectiveness of interventions, taking into account the indirect effects, will lead to better-informed decisions by policymakers.

## Data Availability

All data produced in the present study are available upon reasonable request to the authors

## Declarations Acknowledgments

We are grateful to Dr. Masaru Nagashima for his thoughtful input from his expertise in development economics research.

## Contributions

YKK developed the original concept. YKK and SMM conducted the first literature screening. YKK, WK, CWC, MK, and AKR conducted the full-text reading. YKK drafted the first draft of the manuscript and YKK, WK, CWC, MK, AKR, BNK, MN, JG, and AK contributed to the revisions. All authors reviewed and approved the final manuscript.

