## Supplementary file for "Unraveling the “indirect effects” of interventions against malaria endemicity: A systematic scoping review"

**Supplementary Table 1: Data extraction description**

| Country | The country in which the study was conducted. If the study included more than one country, all countries were listed. If unknown, or if the study was not geographically specific, it was left blank. |
| --- | --- |
| Study type | If a study only collected entomological outcomes (e.g., vector density, sporozoite rate [SR], entomological inoculation rate [EIR]), it is classified as an entomological survey. If a study was a secondary analysis of a previous CRT, it is classified as a re-analysis of previous study. “Others” included analysis of passive surveillance data. |
| Type of malaria parasite | P. falciparum, P. vivax, P. ovale, and P. malaria if specified. |
| Baseline malaria endemicity | Free description of malaria incidence, prevalence, EIR, etc. for malaria transmission intensity in the study area before the intervention was implemented. |
| Separate estimated indirect effect for different conditions or not | Whether the study estimated indirect effects separately for different areas or populations. |
| Pre-specified to measure indirect effect or not | Yes only when the measurement of indirect effects was intended at the research design stage and was explicitly mentioned in the main text or the published protocol; otherwise no. |
| Secondary analysis of previous study or not | Yes if the study used the same data that had already been published previously; otherwise no. |
| Methods of indirect effects estimation | [1] comparison between no treatment in the treatment community and the control group, (1) comparison not conditional on treatment density nor geographical distance, (2) pre-post comparisons among those who did not receive the treatment, [2] Comparison conditional on treatment coverage or geographical distance, (1) comparisons within the control area according to distance to the treatment cluster. (2) comparisons within the treatment area according to the coverage among those who received the treatment. (3) comparisons within the treatment area according to the coverage, including both those who received treatment and those who did not. (4) comparisons within the treatment area according to the coverage among those who did not receive the treatment. (5) comparisons within the control area according to the coverage of the nearest treatment clusters, [3] comparisons conditional on other factors such as the repellent and killing effects of ITNs, pre-erythrocytic or blood-stage vaccine efficacy, endemicity of study area, and the connectedness between different areas. Type [3] only applies to mathematical modeling studies. If one of these did not apply, the respective details were recorded as “Others”. |
| Terminology of indirect effects (Title or abstract) | State any words regarding indirect effects used in the title or the abstract. |
| Terminology of indirect effects (Main text) | State any words regarding indirect effects used in the main text. |
| Positive or negative indirect effect was observed or not | Yes if a statistically significant positive or negative effect was reported, or if there was a visually clear difference between comparators, such as in the simulation results, otherwisen no. |

**Supplementary Table 2: Standardized labels for each term for inconsistencies of words**

| Label | Words |
| --- | --- |
| "community/communal" | community effects, community protection, community-level effects, community-level benefits,  community-level protection, community-level impacts, community-wide effects, community-wide suppression,  community-wide benefits, additional community effect benefits, community impact, communal protection,  communal benefits, beneficial community effect |
| "mass/mass killing" | mass effects, mass community effects, mass killing benefit, the effect of mass mosquito killing |
| "indirect" | indirect effects, indirect protective effects, spatial indirect effects, indirect benefits, indirect protection |
| "herd" | herd effects, herd protection, the secondary effects of herd immunity |
| "spillover" | spillover, spillover effects, spatial spillover effects, spatial spillovers, positive spillovers, positive spatial spillover effects |
| "spatial" | spatial effects, spatial effects, spatial protective effects |
| "area-wide" | area-wide effects |
| "assembly" | assembly effects |
| "population" | population-level effects |
| "group-level" | group-level effects |
| "positive externality" | positive externality |
| "dependent happenings" | dependent happenings |

**Supplementary Table 3: Summary of the included studies in terms of intervention types and indirect effects analysis methods.**

| Intervention types | Methods | N |
| --- | --- | --- |
| ITN/LLIN | [1]-(1): comparison between no treatment in the treatment community and the control group | 2 |
|  | [2]-(1): comparisons within the control area according to distance to the treatment cluster | 5 |
|  | [2]-(2): comparisons within the treatment area according to the coverage among those who received the treatment | 4 |
|  | [2]-(3): comparisons within the treatment area according to the coverage, including both those who received treatment and those who did not | 6 |
|  | [2]-(4): comparisons within the treatment area according to the coverage among those who did not receive the treatment | 7 |
|  | [2]-(5): comparisons within the control area according to the coverage of the nearest treatment clusters | 2 |
|  | [3]: comparisons conditional on the connectedness between different areas | 1 |
|  | Others: reallocating the treatment and control cluster assignments based on the distance to the nearest treatment cluster | 1 |
|  | Others: using counterfactual hypothetical models | 1 |
| House modification | [1]-(1): comparison between no treatment in the treatment community and the control group | 1 |
|  | [2]-(3): comparisons within the treatment area according to the coverage, including both those who received treatment and those who did not | 1 |
|  | [2]-(4): comparisons within the treatment area according to the coverage among those who did not receive the treatment | 1 |
| ITN/LLIN and IRS | [1]-(1): comparison between no treatment in the treatment community and the control group | 1 |
|  | [2]-(2): comparisons within the treatment area according to the coverage among those who received the treatment | 1 |
|  | [2]-(4): comparisons within the treatment area according to the coverage among those who did not receive the treatment | 1 |
|  | [3]: comparisons conditional on the repellent and killing effects on mosquitos | 1 |
| IPT/SMC | [1]-(1): comparison between no treatment in the treatment community and the control group | 2 |
|  | Others: comparison between treatment and control communities with low treatment coverage | 1 |
| MDA | [1]-(2): pre-post comparisons among those who did not receive the treatment | 1 |
|  | [2]-(2): comparisons within the treatment area according to the coverage among those who received the treatment | 1 |
|  | [2]-(3): comparisons within the treatment area according to the coverage, including both those who received treatment and those who did not | 1 |
|  | [2]-(4): comparisons within the treatment area according to the coverage among those who did not receive the treatment | 1 |
|  | [3]: comparisons conditional on the connectedness between different areas and the endemicity of study area. | 1 |
| Malaria Vaccine | [1]-(1): comparison between no treatment in the treatment community and the control group | 1 |
|  | [1]-(2): pre-post comparisons among those who did not receive the treatment | 1 |
|  | [2]-(3): comparisons within the treatment area according to the coverage, including both those who received treatment and those who did not | 1 |
|  | [3]: comparisons conditional on pre-erythrocytic or blood-stage vaccine efficacy | 1 |
| IRS | [1]-(2): pre-post comparisons among those who did not receive the treatment | 1 |
| Untreated bed net | [1]-(2): pre-post comparisons among those who did not receive the treatment | 1 |
